# Volitional deep brain stimulation following brain-computer interface training for Parkinson’s disease

**DOI:** 10.64898/2026.08.12.26350419

**Authors:** Jin-Xiao Zhang, Jiyeon Suh, Pria Daniel, Philip Starr, Jeffrey Herron, Simon Little

## Abstract

Deep brain stimulation (DBS) is transforming from a static therapy toward adaptive systems that adjust stimulation based on neural biomarkers. However, the detection of reliable biomarkers that capture the multi-dimensional nature of complex symptoms is often challenging. Here we demonstrate volitional DBS (vDBS)—a paradigm in which patients use brain-computer interface (BCI) training to learn self-regulation of a neural signal that then controls closed-loop DBS. Two patients with Parkinson’s disease implanted with sensing-enabled neurostimulators completed chronic, at-home BCI training by playing an airplane simulation game. Through training, they were able to effectively down-regulate their cortical beta signal (*p’s* < 1e-10), represented as the real-time position of a plane in the BCI game. Following training, this cortical beta signal served as the input to a closed-loop DBS algorithm. By modulating their beta signal to cross personalized thresholds, patients voluntarily increased or decreased neurostimulation amplitude at will, in the absence of physical movement (*p’s* < 1e-10). This proof-of-principle demonstration establishes that volitional control of intracranial neurostimulation is achievable without the need of an externalized manual controller. BCI-vDBS could potentially be used for a range of neuropsychiatric conditions and brain rehabilitation to support personalized control of neurostimulation.

## INTRODUCTION

Neurological and psychiatric disorders, ranging from Parkinson’s disease (PD) to depression, represent a global health burden with limited treatment efficacy for many patients. In this context, two rapidly advancing neurotechnologies in biomedical engineering, closed-loop neuromodulation and brain-computer interfacing (BCI), have emerged as transformative tools for ameliorating pathological brain states. Integrating closed-loop neuromodulation with BCI technology in therapies could combine advantages from both technologies and create new intervention opportunities for neurological and psychiatric disorders (*1*). However, despite the conceptual synergy, there is a lack of research demonstrations and clinical validation for the integration of these technologies in therapeutic contexts.

Deep Brain Stimulation (DBS) is an established and effective neuromodulation therapy for advanced neurological conditions, especially PD. However, conventional DBS (cDBS) operates with fixed stimulation parameters, which do not adapt to fluctuations in patient symptoms, medication status, or behavioral states. This fixed property of cDBS limits its therapeutic precision and can result in suboptimal control and undesirable side effects. In practice, patients are usually provided with an externalized controller with which they can manually adjust stimulation within safe clinical limits. However, these manual controllers are difficult to operate while experiencing movement symptoms (e.g., tremor, bradykinesia) and almost impossible to implement on a moment-to-moment basis (*2*). To address these limitations, adaptive DBS (aDBS) therapies have emerged, which adjust stimulation in a dynamic manner, usually in response to real-time neural biomarkers (*3*). Clinical studies have shown that aDBS can outperform cDBS in both efficacy and efficiency, and hence, is a promising approach to precise intervention for PD (*4–6*). Nevertheless, the complexity of aDBS programming increases substantially for complex symptom states, such as in patients with fluctuating motor symptoms or heterogeneous non-motor symptoms. These may require more sophisticated, time-intensive programming strategies to optimize therapy and avoid side effects (*7*, *8*).

The success of aDBS hinges on identifying biomarkers that accurately reflect clinical states chronically. In PD, beta oscillatory activity in the cortico–basal ganglia circuit is the most established biomarker (*9*, *10*). Thus, aDBS therapies based on basal ganglia beta have been developed for PD and shown to outperform cDBS in both efficacy and efficiency (*3*, *11–13*). However, this approach faces a number of challenges. The first challenge is that this signal can be confounded by factors such as physical movement, sleep, and the patient’s medication state. This challenge might be greater for psychiatric conditions receiving neurostimulation therapies (*14*, *15*) - such as depression, anxiety, and obsessive-compulsive disorder - where symptoms are more difficult to quantify objectively and clear physiomarkers are lacking (*16*). A further underappreciated challenge is that patients receiving current aDBS approaches may learn to regulate the target biomarker (e.g., beta signal) over time, either implicitly or explicitly, through neurofeedback-like mechanisms. This possibility is grounded in the principle that individuals can learn to alter a changing neural signal when the change is linked to a reward (*17*). In the case of aDBS, symptom alleviation due to neurostimulation change would serve as that reward. This neural learning phenomenon could further complicate biomarker interpretation and utilization. One promising way to address these challenges is to move beyond purely biomarker-reactive stimulation and explore proactive approaches that incorporate patient self-regulation.

The brain is fundamentally a self-regulating system, maintaining homeostasis across cognitive, emotional, and motor domains through continuous feedback and adjustment. In neurological and psychiatric disorders, this capacity for self-regulation becomes disrupted. From this perspective, current aDBS strategies use low-dimensional, externally-derived algorithms to compensate for disrupted neural regulation. A proactive approach that incorporates the patient into this regulatory process could extend therapeutic efficacy. By engaging the patient as an active participant, the entire brain—with its privileged access to internal states, intentions, and context—becomes part of the control system, rather than relying solely on algorithms operating on limited biomarker information. This may be particularly relevant for PD, where patients often know precisely what movement they wish to execute but cannot initiate or execute it satisfactorily. A volitional neurostimulation system that allows patients to bridge this gap through trained self-regulation may more directly address the core phenomenology of the disease than purely reactive approaches.

BCIs have been used for rehabilitative purposes to restore mobility in conditions such as stroke and spinal cord injury (*18*, *19*). During motor imagery training, brain signals from the motor cortex are decoded to provide an external feedback to the patient, in the form of robotic hand movement, electrical stimulation on the targeted skeletal muscle, or visual rewards on a computer screen (*20–24*). For brain disorders stemming from central nervous system pathology (e.g., PD), pairing a BCI with direct neurostimulation may represent a significant advance over traditional rehabilitative BCIs, because the feedback is delivered as intracranial stimulation directly to neural circuits rather than indirectly to peripheral muscles or via the visual system. Direct neurostimulation in a state-dependent manner may engage reinforcement-like mechanisms and facilitate neural plasticity, providing the foundation for an augmented rehabilitation paradigm (*25*, *26*). We envisage that patients could theoretically learn to self-modulate neurostimulation therapy by entering specific optimal neural states that are encoded within the BCI system. With repeated state-dependent stimulation over time, these neural states could be further promoted through selective induction of plasticity and reinforcement.

Here, we implemented a proof-of-principle demonstration of *volitional* DBS (vDBS) in two patients with PD, by integrating DBS with a BCI in a closed-loop system. With this vDBS system, patients voluntarily adjusted their DBS amplitude at will, on a moment-to-moment basis. We first validated previous reports of self-regulation of beta activity with BCI training (*27–30*), and then we showed that through this trained beta self-regulation, patients could directly modulate their neurostimulation amplitude. The clinical aim was to test whether self-modulated neurostimulation is achievable, sustained, has side effects, and to establish a foundation for vDBS systems. It also explicitly tests the possibility of patients learning to regulate the beta signal over time in the naturalistic environment, a target biomarker which has already been used in aDBS therapies for PD.

## RESULTS

We designed an intracranial BCI simulation game that controlled an airplane flight to provide real-time visual feedback of participants’ cortical beta power and guide them to regulate this brain signal via successful scoring in the game (Fig. 1A; Video S1). To evaluate the feasibility of BCI-vDBS, we implemented a two-stage BCI protocol comprising a training phase followed by a testing phase (Fig. 1B). Two right-handed PD patients with chronically implanted sensing-enabled investigational Summit RC+S neurostimulators were recruited from a research clinical cohort (ClinicalTrials.gov registration: NCT03582891) (*6*). In this fully remote, home-based study, they completed all sessions in an ON-medication state (see Methods for protocol details). During the training phase, participants played the BCI game to learn to down-regulate their left cortical beta power (15–20 Hz) without making physical movements. After this ability was established in the training phase, the same cortical beta signal was used in the testing phase as a real-time control input to modulate DBS amplitude dynamically (i.e., vDBS). We assessed patients’ BCI game performance and associated neural activity, as well as pre- and post-game motor performance.

**Fig. 1.**
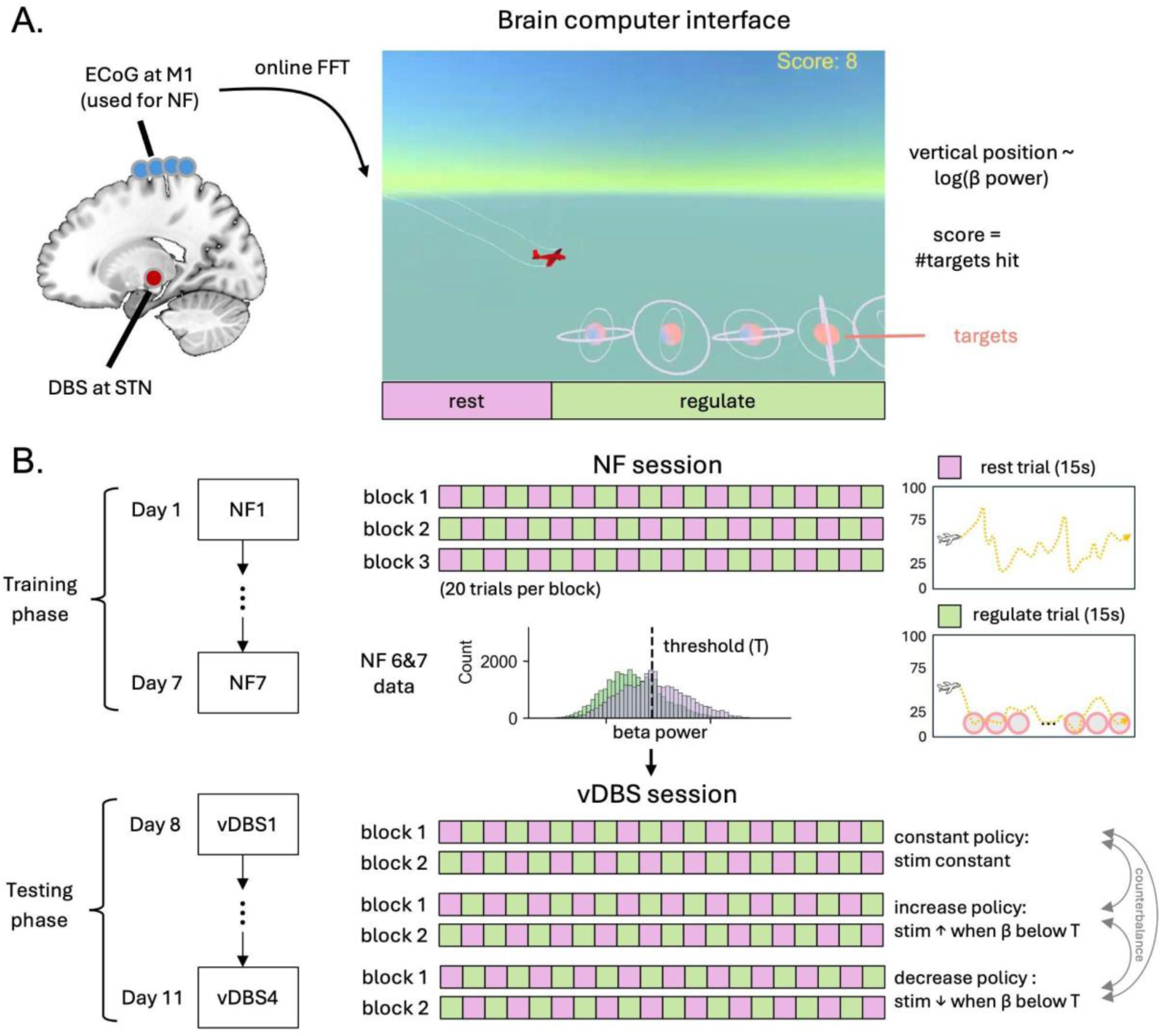
Overview of the brain-computer interface (BCI) study. (A) The BCI airplane simulation game. Two right-handed patients were chronically implanted with bilateral electrocorticography (ECoG) at the sensorimotor cortex and sensing-enabled deep brain stimulation (DBS) lead at the subthalamic nucleus (STN). The vertical position of the plane reflects the log beta power at the left primary motor cortex (M1) channel in real time. The game has ‘rest’ trials and ‘regulate’ trials (see Methods). Participants are required not to make physical movements and are videotaped during the game to check for immobility. (B) Experiment procedure consists of a training phase of 7 neurofeedback (NF) sessions on constant DBS + a testing phase of 4 volitional DBS (vDBS) sessions. Each NF session consists of 3 blocks, with 10 ‘rest’ trials and 10 ‘regulate’ trials in an interleaved order in each block. Data during the last two NF sessions (i.e. NF 6 and 7) were used to determine the best beta threshold (T) for later aDBS tests. Each aDBS session consists of 2 blocks of ‘constant’ policy, 2 blocks of ‘increase’ vDBS policy, and 2 blocks of ‘decrease’ vDBS policy (see Methods). The order of the 3 policies was counterbalanced across vDBS sessions.

### BCI game performance

Across the 7 training sessions, both participants showed performance improvements (i.e., increases in game scores) in the BCI game under constant DBS (Fig. 2A). During the testing session (Fig. 2B), we included additionally an ‘increase’ and a ‘decrease’ DBS policy (see Methods for details) to test game performance under vDBS. Both participants’ performance in the BCI game remained stable across the vDBS policies (Patient 1: *F*(2, 33) = 0.38, *p* = .687; Patient 2: *F*(2, 33) = 0.02, *p* = .977), indicating that the different stimulation conditions did not impair the learned BCI control. Of note, Patient 1 performed their 4th training session approximately 3 hours earlier than their usual time of day, which likely explains the drop in score in this particular session.

**Fig. 2.**
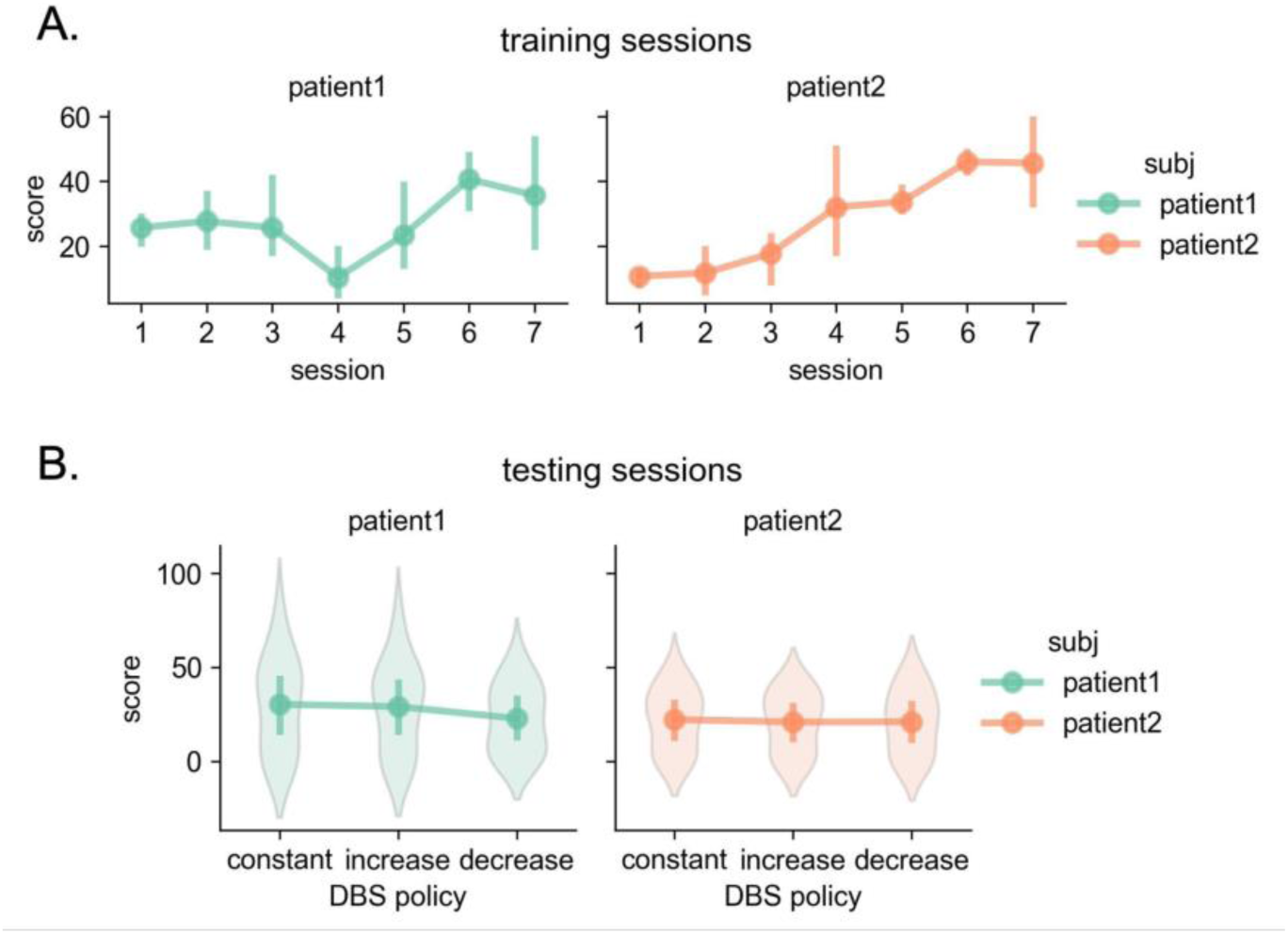
Performance during the BCI game. (A) Participants’ game score improved across training sessions. (B) Participants’ game score across vDBS policies during testing sessions. Their BCI control performance was not negatively impacted by the vDBS algorithm. The score in the game is the number of targets hit by the airplane per block, with a maximum score possible being 150 per block. Error bars represent bootstrapped 95% confidence intervals.

### Successful self-regulation of beta power

During the testing sessions, both participants showed reliable capability for self-modulating their beta power in the ‘regulation’ trials (for training sessions, see Supplementary Fig. S1). Under constant DBS, the trial averaged power reduction in ‘regulation’ trials was observed across a broad beta band (13-30 Hz) in cortical channels (Fig. 3A; Patient 1: *t*(158) = - 7.10, Cohen’s *d* = −1.12, *p* < 1e-10; Patient 2: *t*(158) = −19.49, Cohen’s *d* = −3.08, *p* < 1e-10), which occurred despite the BCI-neurofeedback target being confined to a narrower low beta band. Moreover, despite training on cortical beta, a smaller but significant attenuation in a high-beta power (20-30 Hz) in the subthalamic nucleus (STN) was also evident in both patients, in the presence of subcortical stimulation (Fig. 3B; Patient 1: *t*(158) = −4.43, Cohen’s *d* = −0.70, *p* = 1.78e-5; Patient 2: *t*(158) = −2.14, Cohen’s *d* = −0.34, *p* = 0.034). This supports that learned cortical regulation propagated through the motor network. Similar regulation of beta power was observed under ‘increase’ and ‘decrease’ DBS policies (see Supplementary Fig. S2).

**Fig. 3.**
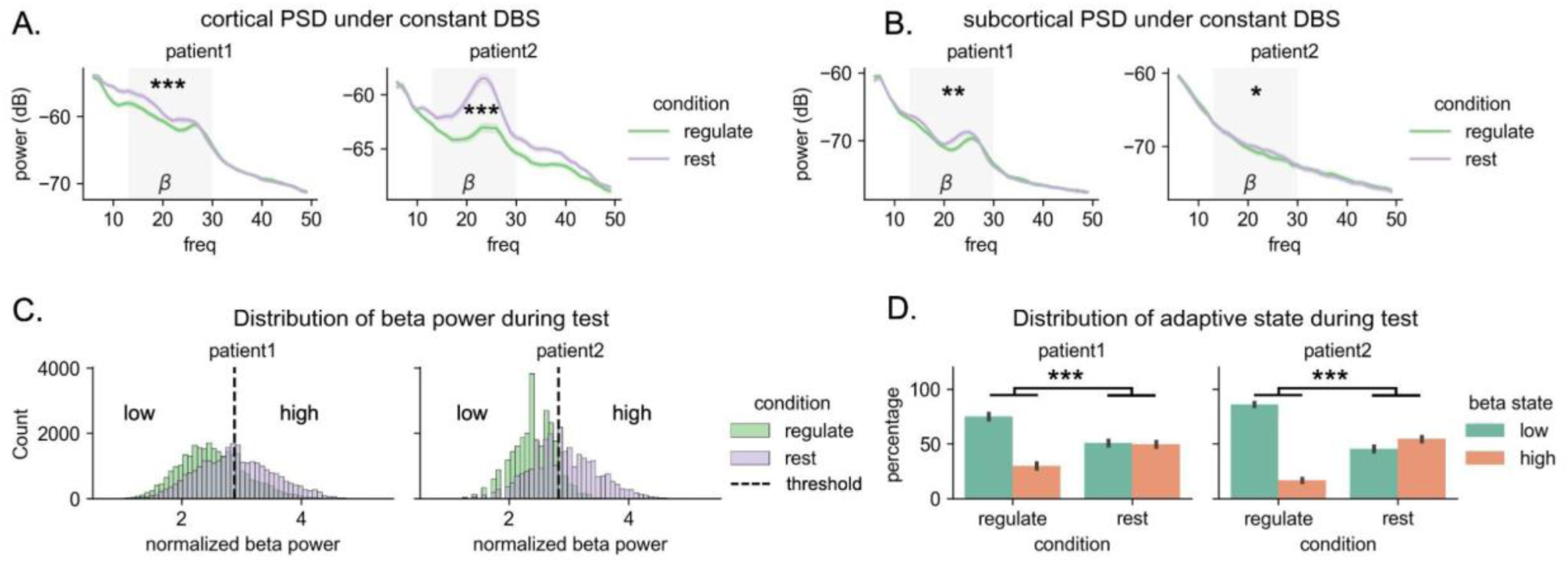
Self-regulation of beta power under constant DBS. (A) Trial averaged cortical power spectral density (PSD) across conditions under constant DBS policy. (B) Trial averaged subcortical power spectral density (PSD) across conditions under constant DBS policy. The grey area represents the beta band (13-30 Hz). Error bars and ribbons represent bootstrapped 95% confidence intervals of the trial averaged values. (C) Distribution separation of real-time cortical beta power (every 0.1 s) between the ‘regulation’ condition and the ‘rest’ condition in an example test session. The dashed vertical line indicates the optimal threshold of beta power to maximize F1 score for each patient. (D) The percentage of time spent in low or high beta state across conditions. Low beta state is when beta power is below the threshold and high beta state is when beta power is above the threshold. Statistical significance: \**p* < .05, \*\**p* < 1e-3, \*\*\**p* < 1e-10. Error bars represent bootstrapped 95% confidence intervals.

Demonstrating moment-to-moment regulation, distinct differences in cortical beta power distributions between the rapid ‘regulation’ and ‘rest’ conditions were evident in the online real-time data for both patients (Fig. 3C; Patient 1: *t*(56082) = −72.99, Cohen’s *d* = −0.62, *p* < 1e-10; Patient 2: *t*(55730) = −120.57, Cohen’s *d* = −1.02, *p* < 1e-10). Based on the data from two most recent training sessions, we set a personalized optimal beta power threshold for each participant during testing sessions (see Methods). With this threshold, the low vs. high beta state percentage within a trial was significantly different between the ‘regulation’ and ‘rest’ conditions, as shown by the condition x beta state interaction (Fig. 3D; Patient 1: *F*(1, 915) = 314.7, *p* < 1e-10, Patient 2: *F*(1, 916) = 1540.7, *p* < 1e-10). This interaction reveals that participants spent most of the time in a low beta state in the ‘regulation’ condition (Patient 1: 75.0%, Patient 2: 86.1%), while they spent much less time in a low beta state in the ‘rest’ condition (Patient 1: 50.8%, Patient 2: 45.6%).

### Volitional control of DBS

Following BCI training on the cortical beta signal, this biomarker was then used as an input signal to the closed-loop DBS to enable volitional neurostimulation adjustments. In the ‘constant’ policy (Fig. 4A), the vDBS control algorithm directed the participants’ stimulation amplitude to stay constant at their regular clinical levels (Patient 1 = 3.0 mA, Patient 2 = 4.0 mA) regardless of their cortical beta state. In the ‘increase’ policy, the vDBS control algorithm was designed to increase the participants’ stimulation amplitude (Patient 1: 3.0 → 3.4 mA, Patient 2: 3.7 → 4.3 mA) when their cortical beta dropped past the personalized threshold. Conversely, in the ‘decrease’ policy, the control algorithm was designed to decrease stimulation amplitude (Patient 1: 3.0 → 2.6 mA, Patient 2: 4.3 → 3.7 mA) when the cortical beta dropped past the threshold.

**Fig. 4.**
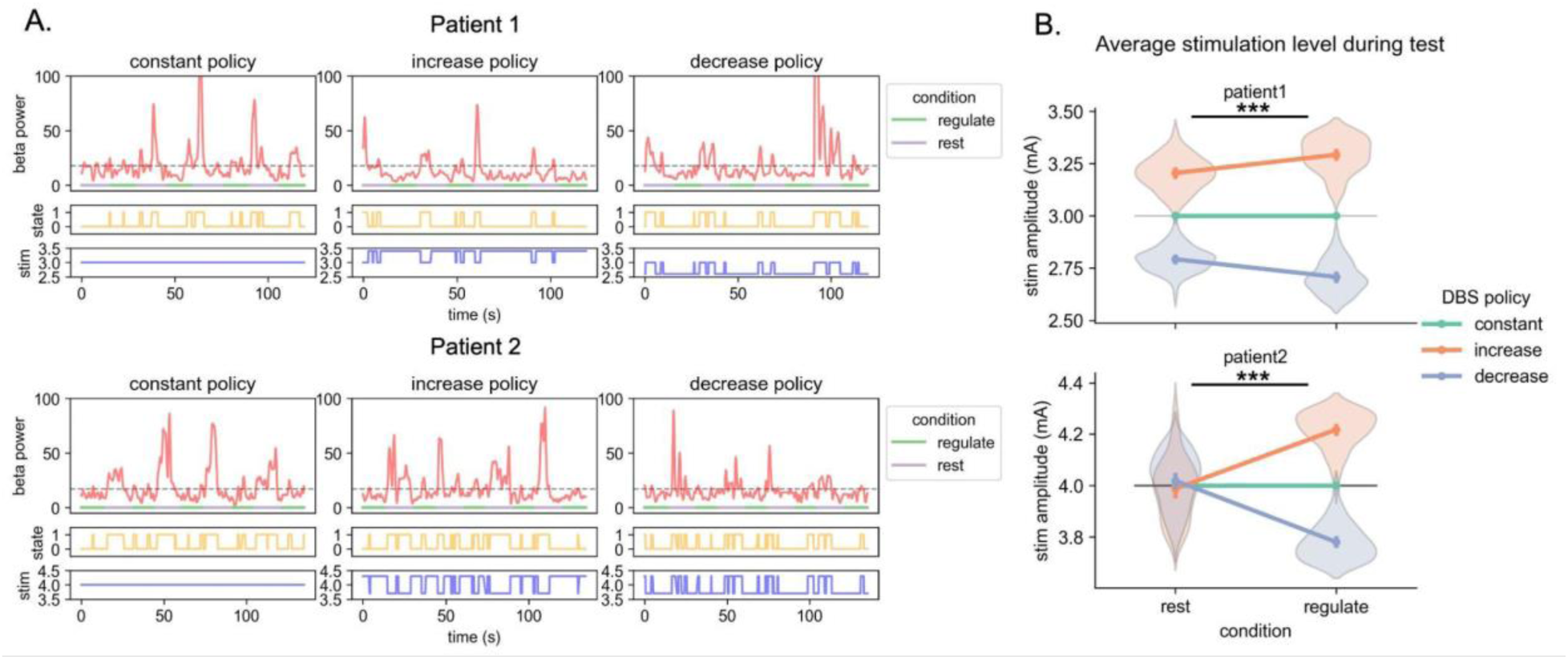
Volitional control of DBS in testing sessions. (A) Temporal dynamics of cortical beta power, its beta state, and stimulation amplitude under the vDBS policies. State 0 indicates a low beta state and state 1 indicates a high beta state. Dashed lines indicate the personalized threshold of beta power. (B) Average stimulation amplitude participants received across trial conditions under the vDBS policies. Significant condition x DBS policy interaction effect was observed in both patients. Statistical significance: \**p* < .05, \*\**p* < 1e-3, \*\*\**p* < 1e-10. Error bars represent bootstrapped 95% confidence intervals.

To quantify the volitional stimulation change, we calculated the trial-level average stimulation amplitude in each trial condition and for each DBS policy. In order to test the effects of condition (rest vs. regulation) x DBS policy (constant, increase, decrease) together, we ran a two-way ANOVA on the average stimulation amplitude. We found a significant condition x DBS policy interactions, Patient 1, *F*(1, 476) = 78.4, *p* < 1e-10, Patient 2, *F*(1, 474) = 366.8, *p* < 1e-10 (Fig. 4B). Unpacking this interaction revealed that, as designed, both patients received significantly greater stimulation in the ‘regulation’ condition than the ‘rest’ condition under the ‘increase’ policy (Patient 1: *t*(159) = 6.88, Cohen’s *d* = 1.09, *p* = 1.3e-10; Patient 2: *t*(158) = 14.62, Cohen’s *d* = 2.31, *p* < 1e-10). Conversely, under the ‘decrease’ policy, they received significantly smaller stimulation in the ‘regulation’ condition than the ‘rest’ condition (Patient 1: *t*(159) = −7.69, Cohen’s *d* = −1.21, *p* < 1e-10; Patient 2: *t*(158) = −16.80, Cohen’s *d* = −2.66, *p* < 1e-10). This indicates that both of the patients voluntarily modulated their DBS stimulation by regulating their cortical beta power in the absence of movements (confirmed through validation of video recordings).

### BCI effects on motor performance

In testing sessions, we assessed the motor performance of the dominant hand before and after BCI game blocks with a key tapping task (see Methods) under the constant DBS policy. We hypothesized a beneficial effect of BCI training so that motor performance would be better post-BCI than pre-BCI. We tested this hypothesis with a paired t-test in each patient. We found a significant effect of BCI training on the tapping speed showing that the patients were faster after BCI training than before (Fig. 5; Patient 1: *t*(39) = 2.52, Cohen’s *d* = 0.40, *p* = .016; Patient 2: *t*(39) = 2.42, Cohen’s *d* = 0.38, *p* = .020), supporting a direct effect of the BCI guided down-regulation of cortical and associated subcortical beta on movement speed.

**Fig. 5.**
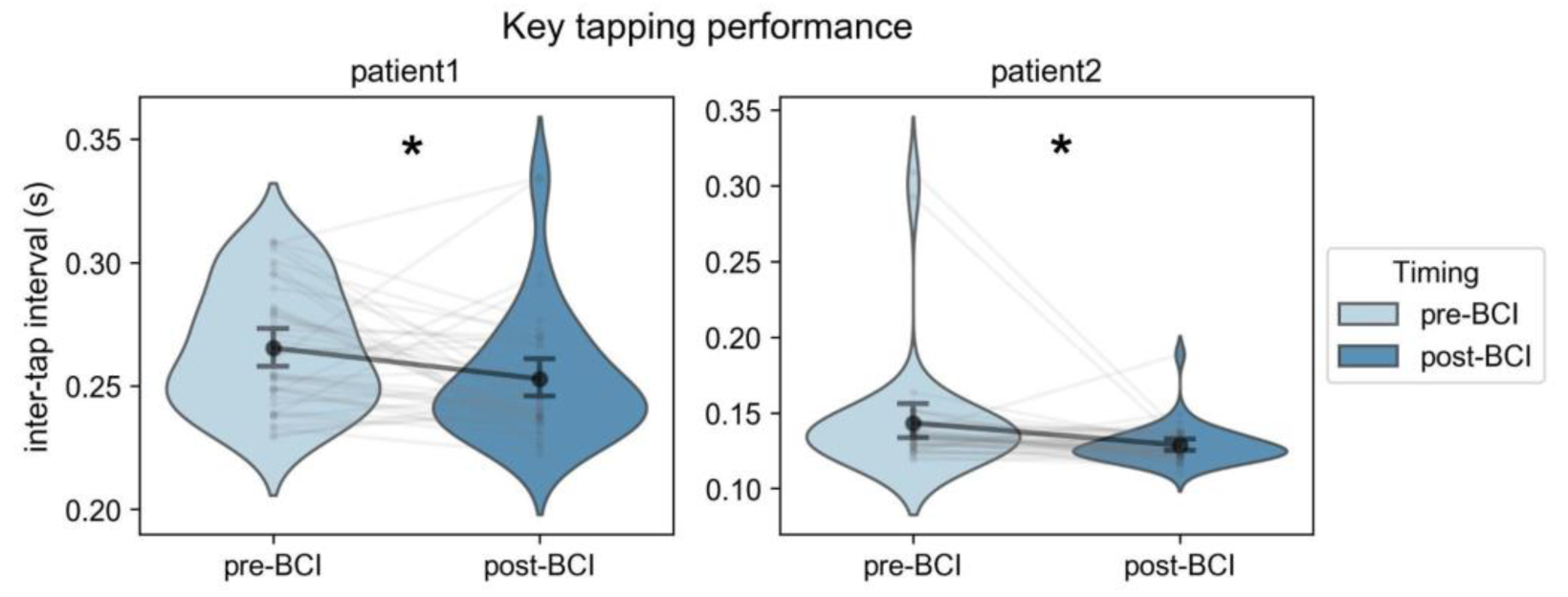
Motor performance in a key tapping task. Participants completed this motor task before and after BCI game blocks during testing sessions. Trial-level tapping speed was calculated as the inter-tap intervals (s) during the key tapping task under constant DBS. Both patients showed improved tapping speed after completing the BCI blocks. Statistical significance: \**p* < .05. Error bars represent bootstrapped 95% confidence intervals.

## DISCUSSION

In two patients with Parkinson’s disease, we demonstrated BCI-based vDBS in a naturalistic home setting. Through BCI training, the patients were able to voluntarily modulate their cortical (and associated subcortical) beta power which was used as an input in a closed-loop DBS system for adjusting neurostimulation amplitude. The stimulation amplitude changes achieved through volitional control were within a clinically relevant range, as the amplitude differences between increase and decrease policies were comparable to those used in conventional aDBS paradigms. The BCI training itself resulted in a speeding of hand movements. This proof-of-principle establishes vDBS as a feasible neurostimulation paradigm with two distinct implications: as a novel therapeutic approach and as an underappreciated factor in existing adaptive neuromodulation systems.

### Therapeutic potential of volitional neurostimulation

The volitional approach potentially addresses a limitation of current aDBS: reliance on simple biomarker driven algorithms with access to only a fraction of the neural information inside the patient’s own brain. In this approach, the patient is recruited as an active contributor to enhance therapeutic control, by leveraging the brain’s unique ability to self-regulate after comparing the current state to a goal state. This may be particularly valuable for conditions where reliable biomarkers remain elusive, including complex motor phenotypes, psychiatric disorders, and heterogeneous symptom presentations. Moreover, by harnessing self-regulation, rather than just external regulation from a closed-loop algorithm, this approach may lead to accumulative gains over time, as hinted here with tapping speed increases from BCI training.

We used a classical neural signal of movement, beta power at primary motor cortex, as the target biomarker in BCI training for proof of principle. One strength of beta power is that it is relatively easy to modulate with BCI training, as demonstrated by prior research (*27–29*). A further advantage is that down-regulation of cortical beta itself was associated with an improvement in motor performance (Fig. 5A), providing an additive therapeutic effect. However, this cortical beta signal could be confounded by physical movements and medication usage in real life. When making a physical movement, cortical beta oscillations are likely to desynchronize (i.e., a decrease in power) (*31*). This property makes our vDBS under the ‘increase’ policy have parallels to the recently developed, albeit more sophisticated classifier based movement-responsive DBS (*4*). Notably, learned cortical beta regulation was not confined to the cortical recording site but propagated to STN (Fig. 3B). This cortico-subthalamic propagation suggests that volitional regulation engages the motor circuit at the level of the therapeutic target, rather than representing a purely cortical phenomenon. In PD, beta activities (particularly subcortically) are related to the patient’s dopaminergic medication levels and motor symptoms (*32*, *33*), so that the beta baseline level fluctuates along with the medication cycle and beta bursts occur more frequently when off-medication. Whether other neural signals could be a robustly modulable biomarker with fewer confounds warrants further research.

Besides movement disorders, the volitional neuromodulation methodology presents a potentially powerful extension of current rehabilitative BCIs. A cornerstone of effective motor rehabilitation, particularly post-stroke, is the induction of meaningful neuroplasticity within the lesioned motor network. Current rehabilitative BCIs typically rely on peripheral feedback—such as functional electrical stimulation, robotic actuation, or visual rewards—to reinforce the patient’s motor imagery (*18*, *19*). In contrast, our system couples visual BCI with electrical neurostimulation applied directly to the motor circuit. This direct intervention offers a fundamentally more precise, and potentially more potent mechanism for immediately reinforcing the activity of behaviorally active motor circuits (*26*). Furthermore, this dual feedback mechanism, which provides both the visual BCI signal and the therapeutic effect of contingent neurostimulation, could be leveraged to re-establish functional neural pathways with potentially greater efficacy than peripheral approaches alone. Therefore, the BCI-vDBS paradigm developed here may provide a potential pathway for accelerating motor recovery and enhancing functional gains after brain injuries and stroke.

### Implications for adaptive DBS therapies

Despite the opportunities created by volitional neurostimulation, our findings also suggest a potential caveat for existing aDBS therapies and those that are being developed, notably for aDBS policies to treat neurological diseases. We showed that the regulating capability of cortical and subcortical beta power is learnable, and this is potentially true for other brain signals and anatomical targets as well (*27*, *28*). Theoretically, if patients under a current beta-based adaptive DBS policy either like or dislike (even implicitly) the effects of their neurostimulation, they could learn to modulate their stimulation amplitudes by controlling their own brain signals. Observation and interview of our two patients revealed that they shifted their beta-regulating strategy from explicit to more implicit strategies during our BCI training protocol. This phenomenon has been observed in other BCI training paradigms in general, which underlines the possibility of implicit learning. When this implicit learning is strong enough, patients receiving beta-based aDBS may learn to modulate stimulation even at the motor preparation stage due to the predicted movement need. Acknowledging this potential caveat in current beta-based aDBS therapies, researchers and clinicians will need to monitor the possibility of self-modulating beta signals over time and design systems to leverage this while preventing patients from harnessing this potentiality to their detriment.

### Practical considerations

To transform the concept of volitional neurostimulation into a viable therapy, it is important to explore its practicality in clinical scenarios. A primary challenge lies in assessing whether requiring patients to control their own stimulation through mental effort would be distracting and effortful. Note, the neural system learns to regulate many processes implicitly and autonomously (walking, talking, thinking), therefore it is possible that as patients move from explicit to implicit control, the effort of volitional neuromodulation would decrease and may be even eliminated. A second challenge concerns the identification of a target neural signal or a combination of neural signals without interference from normative activities. For instance, beta signals are responsive to movements (*31*) and low-frequency signals (delta and theta) are responsive to sleep and mood (*34*, *35*), making them potentially suboptimal targets. Using an orthogonal signal to movement or sleep for volitional neuromodulation might require longer and more sophisticated training, but may be more robust in the long term. A third challenge concerns whether patients can identify optimal time points to self-regulate their neurostimulation. In the current study, patients were explicitly cued to regulate their neural activity and thereby modulate their neurostimulation. Applying volitional neurostimulation at a suboptimal timing may lead to unexpected results. For instance, PD patients may incorrectly identify medication-off states while their subcortical beta is in fact not high (*36*), which might result in “self-inflicted dyskinesia." A fourth challenge will depend on how long patients can sustain volitional control of BCI-trained brain signals without a reminder training. We anecdotally observed that our two patients were still able to perform cortical beta-regulation after a one-month gap but future studies are required to test the long-term practicality of volitional neurostimulation. In clinical practice, a hybrid system combining reactive aDBS with a secondary volitional mechanism may be optimal.

### Limitations

This study has several limitations. First, this proof-of-principle demonstration involved only two patients. While we showed successful and statistically robust vDBS in both patients, studies with larger cohorts are needed to examine individual heterogeneity in vDBS implementation and explore other potential neural signals for vDBS. Second, although participants were videotaped throughout and instructed not to make physical movements, and post-hoc review of video recordings confirmed the absence of overt movements during regulation trials, we did not use continuous electromyography (EMG) to rule out subthreshold muscle activation. Future studies should incorporate EMG or accelerometry to provide physiological confirmation of movement absence during volitional regulation. Third, we tested the participants’ motor performance before and after the BCI training blocks instead of amidst the BCI training trials. This made us unable to test the effect of BCI guided beta-regulation directly. We did this in order to avoid movement-based disruptions to the target beta signal during the game. Future studies with larger cohorts should systematically examine whether vDBS-driven amplitude changes produce measurable motor effects independent of total stimulation dose, and whether cumulative BCI training leads to sustained motor improvements.

## Conclusion

We establish that patients can learn to volitionally control intracranial neurostimulation through BCI trained self-regulation of neural activity (i.e. volitional DBS). This vDBS paradigm offers a potentially new approach for personalized, patient-driven neuromodulation and highlights the importance of considering learned physiomarker regulation in adaptive neurostimulation therapies. This vDBS approach could be applied to a range of neurological disorders and psychiatric conditions to support personalized control of neurostimulation and augmented rehabilitation.

## MATERIALS AND METHODS

### Participants

Between July and December 2024, we recruited two right-handed PD patients (Table 1) implanted with the Medtronic Summit RC+S investigational neuromodulation system with DBS electrodes at the subthalamic nucleus (STN) and an electrocorticography (ECoG) strip placed over the sensorimotor cortex. The patients had previously participated in a separate clinical trial of chronic multi-site brain recording (ClinicalTrials.gov registration: NCT03582891). They received constant bilateral DBS as an ongoing clinical treatment and were therapeutically optimized by an experienced movement disorders neurologist. Patients’ motor symptoms prior to DBS implantation were assessed using the MDS-UPDRS-III scale. Informed written consent was provided by the patients, and the protocol was approved by the UCSF Institutional Review Board.

**Table 1.**
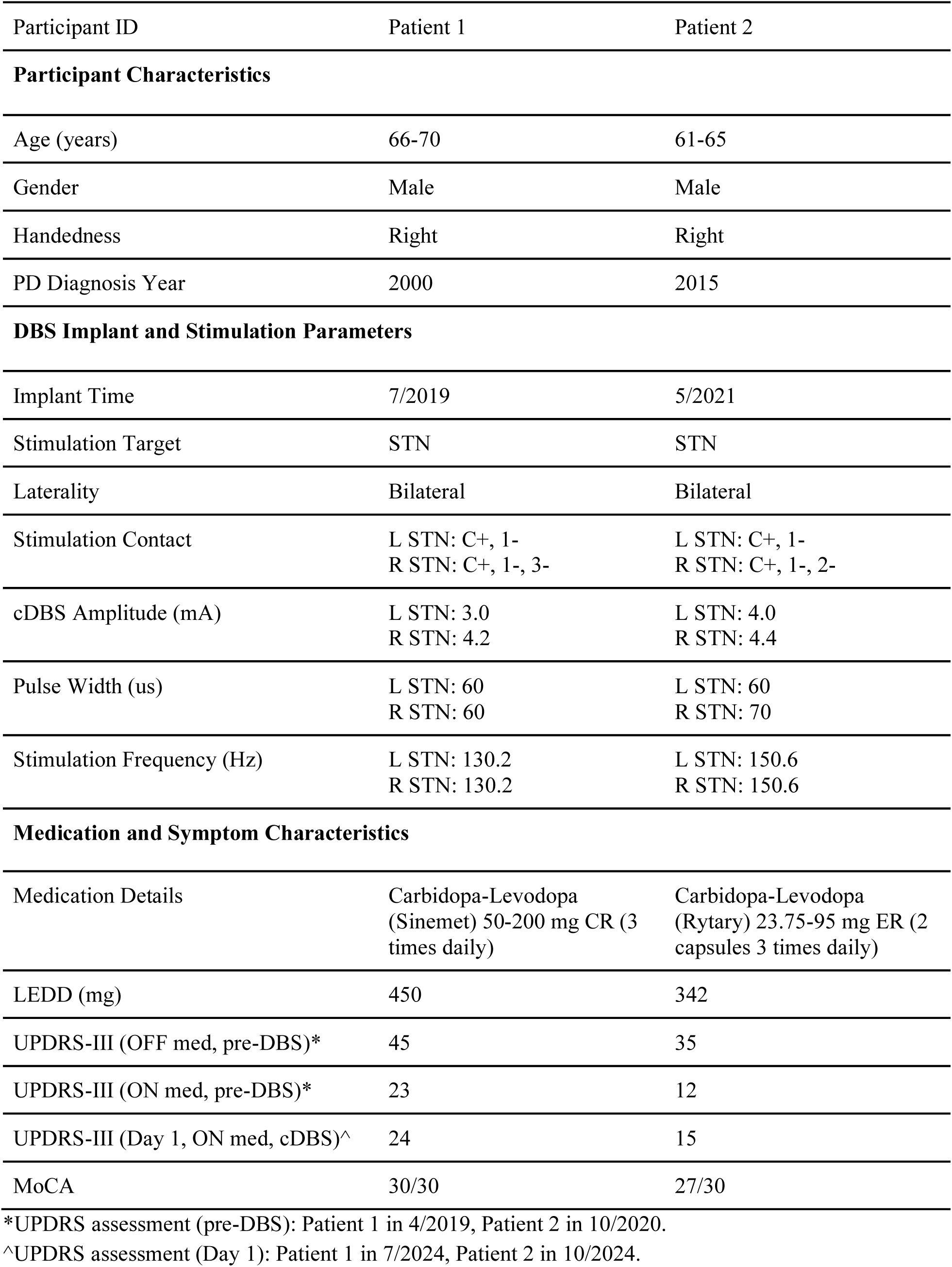
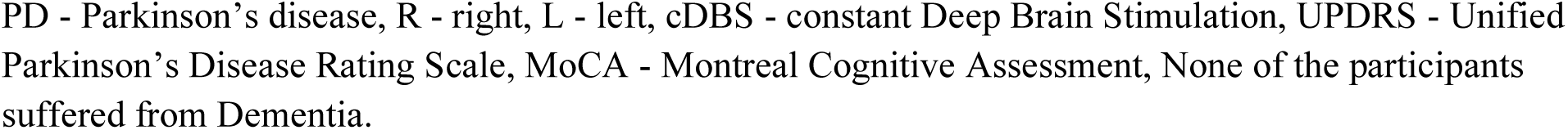
Participants demographics, clinical characteristics and stimulation settings.

| Participant ID | Patient 1 | Patient 2 |
| --- | --- | --- |
| **Participant Characteristics** |  |  |
| Age (years) | 66-70 | 61-65 |
| Gender | Male | Male |
| Handedness | Right | Right |
| PD Diagnosis Year | 2000 | 2015 |
| **DBS Implant and Stimulation Parameters** |  |  |
| Implant Time | 7/2019 | 5/2021 |
| Stimulation Target | STN | STN |
| Laterality | Bilateral | Bilateral |
| Stimulation Contact | L STN: C+, 1-<br>R STN: C+, 1-, 3- | L STN: C+, 1-<br>R STN: C+, 1-, 2- |
| cDBS Amplitude (mA) | L STN: 3.0<br>R STN: 4.2 | L STN: 4.0<br>R STN: 4.4 |
| Pulse Width (us) | L STN: 60<br>R STN: 60 | L STN: 60<br>R STN: 70 |
| Stimulation Frequency (Hz) | L STN: 130.2<br>R STN: 130.2 | L STN: 150.6<br>R STN: 150.6 |
| **Medication and Symptom Characteristics** |  |  |
| Medication Details | Carbidopa-Levodopa<br>(Sinemet) 50-200 mg CR (3<br>times daily) | Carbidopa-Levodopa<br>(Rytary) 23.75-95 mg ER (2<br>capsules 3 times daily) |
| LEDD (mg) | 450 | 342 |
| UPDRS-III (OFF med, pre-DBS)* | 45 | 35 |
| UPDRS-III (ON med, pre-DBS)* | 23 | 12 |
| UPDRS-III (Day 1, ON med, cDBS)^ | 24 | 15 |
| MoCA | 30/30 | 27/30 |

\*UPDRS assessment (pre-DBS): Patient 1 in 4/2019, Patient 2 in 10/2020.
^UPDRS assessment (Day 1): Patient 1 in 7/2024, Patient 2 in 10/2024.
PD - Parkinson's disease, R - right, L - left, cDBS - constant Deep Brain Stimulation, UPDRS - Unified Parkinson's Disease Rating Scale, MoCA - Montreal Cognitive Assessment, None of the participants suffered from Dementia.

### Brain-computer interface

We implemented a BCI training game in which participants fly an airplane icon to hit targets, controlled by real-time intracranial brain signals (Fig. 1A, Video S1). The game was developed in the Unity (https://unity.com/) video game engine using a combination of purchased art assets and custom software development. During gameplay, the plane flies horizontally at a constant speed (maintaining a fixed horizontal position on the screen with the background moving from right to left). The vertical position of the plane was set to be controlled by a Summit RC+S embedded computation of an intracranial signal, here the beta power of the patient’s left primary motor cortex (contralateral to their dominant hand) streamed to the task computer (Microsoft Surface) using the Summit API. In Patient 1, the vertical position of the plane was set to be proportional to the neural signal. In Patient 2, the vertical position of the plane was set to be inversely proportional to the neural signal to counterbalance across patients.

### Neural data streaming

During the BCI game, intracranial time-domain signals of the participants’ left hemisphere (cortical + STN channels) were recorded with a sampling rate of 250 Hz. The beta power (15-20 Hz) of the primary motor cortex (contact 8-10) was calculated online in the embedded device by Fast Fourier Transform (FFT). This beta power was log-transformed to be used for visual feedback in the BCI airplane simulation game. Through piloting, we calibrated this signal to be close-to-normally distributed on a scale of 0-100, mapping to the plane position on the screen (0 = bottom of screen, 100 = top of screen). The online time frequency deconstruction (i.e. FFT) was performed with a window size of 1024 points and an interval of 100 ms. The resulting beta power was streamed to a gRPC-enabled microservice (Open Mind Neuromodulation Interface, https://github.com/openmind-consortium/OmniSummitMicroservice-PublicRelease) on the task PC (*37*), which relayed the control signal to the BCI game in Unity where it was mapped to the airplane vertical position.

### Procedures

Participants went through a protocol of 11 total sessions consisting of 7 training sessions and 4 testing sessions, conducted fully remotely in their homes, on different days (Fig. 1B). On each day, participants started the session one hour after ingestion of the standard PD Levodopa medication. Researchers interacted with the participants remotely through video telemetry, operated the task computer via remote desktop, and updated stimulation programming settings remotely. On Days 1-7 (training sessions), they were trained on down-regulating their cortical beta activity by playing the plane simulation BCI game with visual neurofeedback. At the beginning of the experiment protocol (i.e., Day 1), a neurologist performed a baseline motor examination of the participants through video telemetry using the MDS-UPDRS-III scale and the video recordings were scored by a further neurologist blinded to experiment conditions (Table 1). On Days 8-11 (testing sessions), participants played the BCI game under vDBS (see details below). They were instructed not to make physical movements or contract their musculature throughout the BCI game and were digitally video recorded throughout.

During training sessions, participants played the BCI game under constant DBS at their regular, clinically optimized, settings. There were two trial conditions during the game: ‘rest’ trials and ‘regulate’ trials. During the rest trials, participants were instructed to relax and observe the natural fluctuation of the plane (related to natural cortical beta fluctuations) but not attempt to modulate the plane’s position. During the regulation trials, there were spherical targets that appeared at either the top or bottom 25% of the screen height (Patient 1 at top, Patient 2 at bottom). Compared to the rest condition, participants needed to down-regulate their cortical beta power in order to hit the spherical targets to score. The participants’ goal was to hit as many targets as possible, as indicated by a cumulative score at the top right of the screen. Within each game block, there were interleaved 10 rest trials + 10 regulation trials, with the order counterbalanced across blocks (i.e., rest-regulation or regulation-rest). Participants completed 3 blocks of the game (20 trials per block, 15s per trial, 60s break between blocks). The total session length was approximately 20 minutes. Before and after the BCI training game, participants completed a key tapping task for motor assessment.

During vDBS testing sessions, participants played the same BCI game under three vDBS policies: 1) an ‘increase’ policy, 2) a ‘decrease’ policy, and 3) a ‘constant’ policy. Participants completed two blocks of the game under each policy. Under the ‘increase’ policy, the stimulation amplitude increases when the target brain signal (i.e., cortical beta power) falls below the pre-set threshold. Under the ‘decrease’ policy, the stimulation amplitude decreases when the target brain signal falls below the pre-set threshold. Under the ‘constant’ policy, the stimulation amplitude remains constant at the clinical level. The temporal order of the three DBS policies was counterbalanced across sessions and patients were blinded to stimulation conditions. Under each vDBS policy, participants also completed the same key tapping task before and after the corresponding game blocks.

### Adaptive deep brain stimulation

We used the data from the last two training sessions to determine the beta power threshold for vDBS. The threshold was determined to maximize the differentiation between rest vs. regulation trials during the BCI game by maximizing the F1 score. F1 score was chosen as the threshold optimization criterion because it balances sensitivity and specificity for detecting regulation vs. rest states, which is important given that both false positives (unnecessary stimulation changes) and false negatives (missed regulation attempts) have clinical consequences.

The control algorithm was set up so that when the target brain signal is below the pre-set threshold, it is defined as being in ‘state0’. When the target brain signal is above the pre-set threshold, it is defined as being in ‘state1’. Across all vDBS policies, the overall upper limit of the stimulation amplitude was tested with the participants and set at a maximal level without introducing distraction or discomfort. The overall lower limit of stimulation was set to be symmetrical to the upper limit in reference to the participant’s clinical level. We tested two different sets of adaptive policies in the two participants. In Patient 1, the lower level of stimulation amplitude in the ‘increase’ policy and the upper level in the ‘decrease’ policy were set the same as the clinical stimulation level (i.e., constant policy: stim=3.0 mA; increase policy: state0 = 3.4 mA, state1 = 3.0 mA; decrease policy: state0 = 2.6 mA, state1 = 3.0 mA). In Patient 2, the lower and upper levels within the ‘increase’ and ‘decrease’ policies were set symmetrically in reference to the clinical level (i.e., constant policy: stim=4.0 mA; increase policy: state0 = 4.3 mA, state1 = 3.7 mA; decrease policy: state0 = 3.7 mA, state1 = 4.3 mA). The ramping rate of stimulation change was tested between the lower and upper stimulation levels and set to the maximum within patient tolerance (8.67 mA/s) without inducing side effects (e.g., paresthesias).

### Motor performance assessment

Motor performance was assessed with a key tapping task on the same task computer. In a trial, patients tapped the left and right arrow keys alternately with their index and middle finger of the dominant hand for 10 times as fast as they could. They completed 10 trials during each assessment. Rapid alternating movement speed was quantified as the average inter-tap interval (s) during each trial.

### Data analysis

Intracranial neural data were first preprocessed with Openmind ProcessRCS (https://github.com/openmind-consortium/Analysis-rcs-data) in Matlab R2022a. All other data analysis was performed in Python 3.11.4. Power spectral density was computed with Welch’s method (psd_array_welch in MNE time-frequency) with a Hamming window, 250-point segments (1s sample), and 50% overlap. ANOVA models (Type II) were performed using ‘statsmodels’. Following significant omnibus ANOVA interaction effects, planned pairwise contrasts were conducted without correction for multiple comparisons, consistent with the Fisher LSD approach for planned, hypothesis-driven comparisons (*38*). Independent and paired t-tests were performed using ‘scipy’. Data were visualized with seaborn and matplotlib packages. All error bars/ribbons in plots represent 95% confidence intervals bootstrapped for 1000 times.

## Data Availability

All code and data supporting the statistical analyses of this study are available at https://github.com/tepzhang/volitional_DBS. Any additional requests for materials can be directed to, and will be fulfilled by, the corresponding authors. The raw (identifiable) data from participants are privacy-protected.

https://drive.google.com/file/d/1R4cybpNEjWfQo2Y0tK6O8WWJLfu5Fmm0/view?usp=sharing

https://github.com/tepzhang/volitional_DBS

## Acknowledgement

We thank Kei Landin and Tim Denison for early support on the Unity game development. We thank Elena Ubeda Matzilevich, Amelia Hahn, and Jiaang Yao for their help with running the experiments. We thank Lucia Ricciardi and Victoria Chang for running MDS-UPDRS exams. We thank Gadi Maayan Eshed for scoring the recordings and providing critical feedback to the manuscript. We express our gratitude to the participants for their valuable contributions to this study.

## Author contributions

Conceptualization: JXZ, JH, SL

Methodology: JXZ, JS, PD, JH

Software: JXZ, JH

Investigation: JXZ, JS

Formal Analysis: JXZ, JS

Visualization: JXZ, JS

Resources: PS, SL

Funding acquisition: SL

Supervision: PS, JH, SL

Writing – original draft: JXZ, JS, SL

Writing – review & editing: JXZ, JS, PD, PS, JH, SL

## Supplementary Materials

### Successful volitional regulation of beta power

Across the training sessions, participants showed improvements in self-modulating beta power in the ‘regulation’ trails compared to the ‘rest’ trials. This training effect applies to both cortical (Fig. S1A) and subcortical (Fig. S1B, particularly Patient 1) channels.

**Fig. S1.**
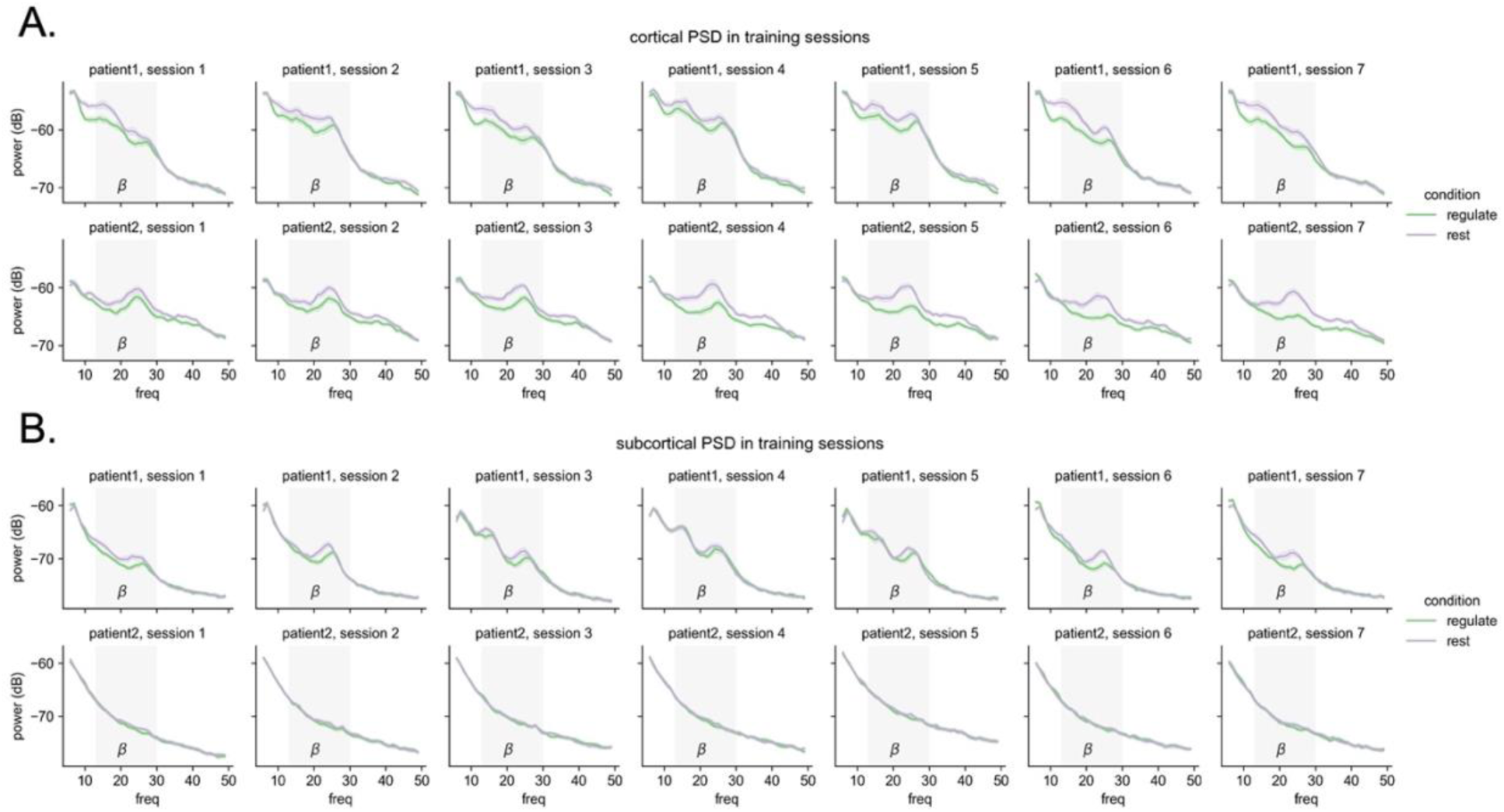
Self-regulation of beta power during training sessions. (A) Trial averaged cortical power spectral density (PSD) during training sessions. (B) Trial averaged subcortical PSD during training sessions. All training sessions were conducted under constant DBS policy. The grey area represents the beta band (13-30 Hz). Error bars and ribbons represent bootstrapped 95% confidence intervals of the trial averaged values.

In order to test the effects of condition (rest vs. regulation) x DBS policy (constant, increase, decrease) together, we ran a two-way ANOVA on the target cortical beta power in the testing sessions. A main effect of condition indicated that the beta power was significantly lower in the ‘regulation’ trial compared to the ‘rest’ trials across DBS policies (Fig. S2A; Patient 1: *F*(1, 474) = 212.9, *p* < 1e-10; Patient 2: *F*(1, 474) = 737.7, *p* < 1e-10). In addition, we found a significant condition x DBS policy interaction on the beta power in Patient 1, *F*(2, 474) = 4.16, *p* = .016, supporting that the beta power regulation capability was stronger in ‘constant’ policy (Cohen’s *d* = 1.67) compared to ‘increase’ (Cohen’s *d* = 1.17) or ‘decrease’ policies (Cohen’s *d* = 1.18). This interaction was not significant in Patient 2, *F*(2, 474) = 1.58, *p* = .207. Furthermore, we examined the temporal dynamics of cortical beta in each trial and calculated the time taken from the trial onset for it to drop to the threshold. Consistently, a condition x DBS policy ANOVA revealed a main effect of condition that patients were much faster to reach the beta threshold in ‘regulation’ trials than in ‘rest’ trials (Fig. S2B; Patient 1: *F*(1, 473) = 61.9, *p* < 1e-10; Patient 2: *F*(1, 474) = 42.4, *p* = 1.9e-10). Additionally in Patient 2, there was a significant condition x DBS policy interaction, *F*(2, 474) = 5.58, *p* = .004, suggesting that this regulation vs. rest condition effect was stronger in ‘constant’ policy (Cohen’s *d* = 0.78) than ‘increase’ (Cohen’s *d* = 0.41) and ‘decrease’ policies (Cohen’s *d* = 0.53).

**Fig. S2.**
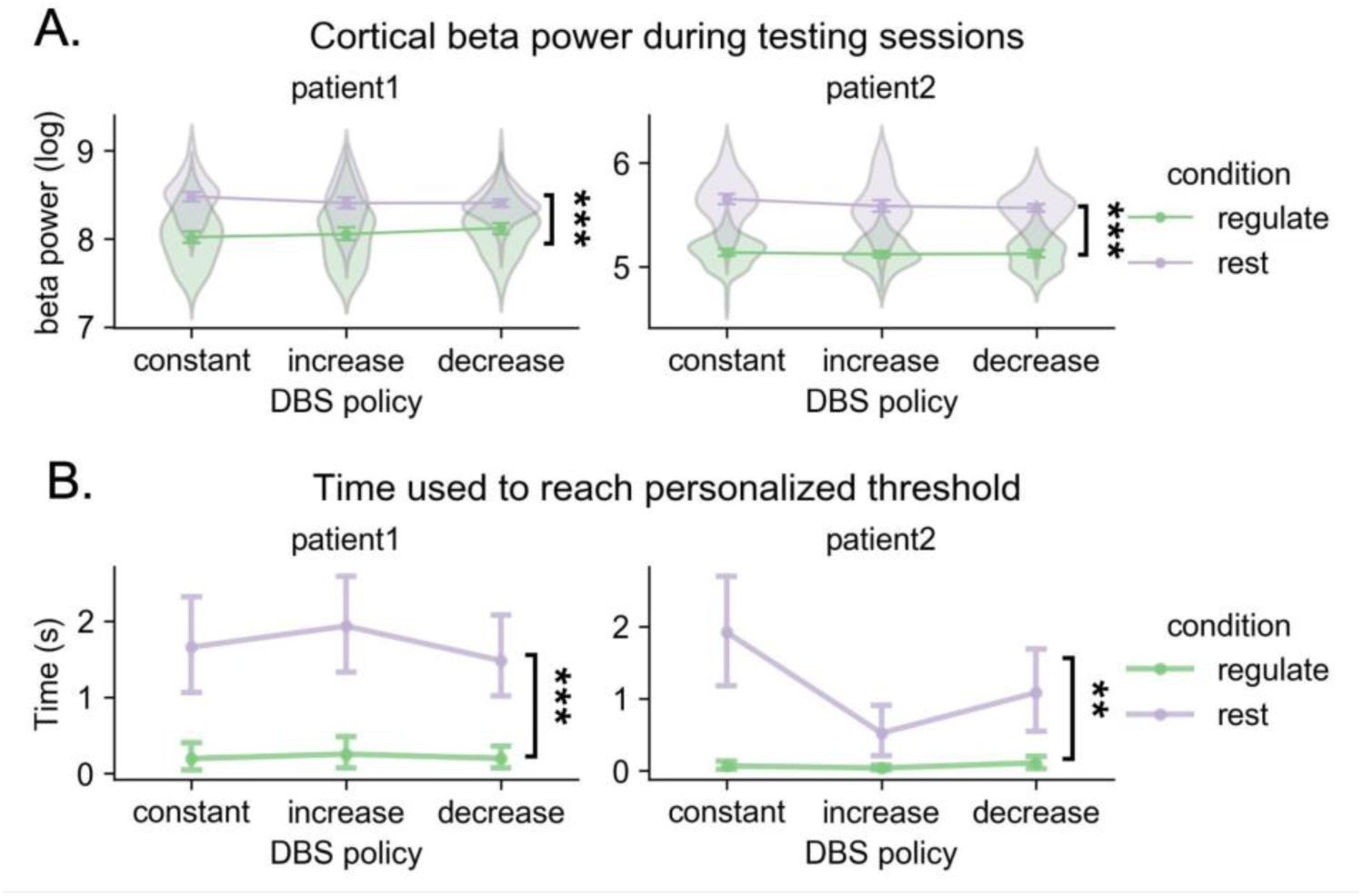
Self-regulation of cortical beta power during testing sessions under vDBS. (A) Trial-level average cortical beta power in the ‘regulation’ and ‘rest’ conditions. Beta power in the ‘regulation’ condition was significantly lower than in the ‘rest’ condition. (B) Time taken for cortical beta to reach the personalized threshold in the ‘regulation’ and ‘rest’ trials. The time used in the ‘regulation’ trials was significantly lower than in the ‘rest’ trials. Statistical significance: \**p* < .05, \*\**p* < 1e-3, \*\*\**p* < 1e-10. Error bars represent bootstrapped 95% confidence intervals.

**Video S1**: demonstration of volitional DBS (link)

